# ‘Trained immunity’ from *Mycobacterium spp*. exposure or BCG vaccination and COVID-19 outcomes

**DOI:** 10.1101/2020.07.11.20151308

**Authors:** Samer Singh, Rajendra P. Maurya, Rakesh K. Singh

## Abstract

Protective variables for COVID-19 are unknown. ‘Trained immunity’ of the populace as a result of BCG immunization policy implementation and coverage had been suggested to be one of the factors responsible for the differential impact of COVID-19 on different countries. Several trials are underway to evaluate the potential protective role of BCG vaccination in COVID-19. However, the lack of clarity on the use of appropriate controls concerning the measures of ‘trained immunity’ or the heterologous cell-mediated immunity conferred by BCG vaccination has been a cause of concern leading to more confusion as exemplified by a recently concluded trial in Israel that failed to find any protective correlation with regard to BCG vaccination. Whereas, when we analyze the COVID-19 data of European countries without any regard for BCG vaccination policy but with similar age distribution, comparable confounding variables, and the stage of the pandemic, the prevalence of tuberculin immunoreactivity - a measure of cell-mediated immunity persistence as a result of *Mycobacterium spp*. (including BCG vaccine) exposure of the populations, is found consistently negatively correlated with COVID-19 infections and mortality per million population, at all the time points evaluated. We propose that on-going and future studies evaluating the effect of BCG vaccination on COVID-19 outcomes may actively consider, if not already, the inclusion of controls for underlying ‘trained immunity’ and heterologous cell-mediated immunity prevalence that may be pre-existing or resulting from the intervention (e.g., BCG vaccine) in such trials to arrive at more dependable conclusions concerning their potential benefit.

## INTRODUCTION

Bacille Calmette-Guérin or BCG vaccination policy implementation by countries and its coverage had been proposed to lower the COVID-19 cases and mortality [REFs in 1-4]. The BCG vaccination given to children and adults has been known to protect them from many unrelated pathogens and diseases [REFs in 1,3, 5-10] supposedly through the generation of ‘trained immunity’ and heterologous cell-mediated immunity (e.g., Th1 and Th17) that last for a variable period [1,5,9-12]. However, the confounding variables among the countries for COVID-19 such as the stage of the outbreak, population age distribution, health infrastructure, management practices, the testing/screening, and reporting guidelines, etc. make the comparisons and conclusions drawn about the protective role of BCG vaccination tenuous [REFs in 2-4]. Notwithstanding, multiple trials are planned and ongoing to assess the potential protective effect of BCG vaccination on COVID-19 incidence and severity [REFs in 1,13]. In a recently concluded study reported in JAMA by Hamiel and colleagues, the individuals vaccinated at birth (born between 1979-1981) were found to be indistinguishable from those not vaccinated at birth (born between 1983-1985) with regard to COVID-19 incidence and severity, negating any protective effect drawn in adulthood from BCG vaccination at birth [14]. It would have been expected from the known complete waning of the neonatal period BCG vaccination imparted cell-mediated immunity within the first few years (<5) after birth, reaching a frequency observed for the reference population due to environmental mycobacteria exposure [REFs in 15,16]. For individuals vaccinated at later stages, the loss is slower and dependent upon other variables as well but estimated to reach background reference population levels with an 8% annual decrease [16]. However, two refined epidemiological analyses that have come up during the review of current commentary [3,4] have again failed in their analysis to confirm the null hypothesis that BCG vaccination is not associated with COVID-19 mortality even after making attempts to minimize the effect of the confounding variables that may have plagued previously observed negative correlation between BCG vaccination policy and COVID-19 incidences and outcomes by adjusting for different parameters, rather these studies suggest that BCG vaccination policy seem to have a protective role as suggested by previous relatively crude analysis [REFs in 2-4]. However, caution needs to be exercised in looking too much into the analysis as it is from an earlier phase of the epidemic when nations clubbed together for the analysis have not reached a similar stage of the epidemic and the possibility of the presence of other potential confounders (see discussion later).

Here we may be reminded of that in the current scenario our primary question is ‘not’ whether BCG vaccination once performed maybe somehow correlated or covarying (not necessarily biologically relevant) with lower incidence and mortality observed in populations and if it can be transformed in a way to model the available data rather the question is a simpler one ‘whether BCG vaccination or any other intervention can potentially reduce/prevent the COVID-19 incidence and mortality in populations’.

Since the delayed cell-mediated immune response, historically measured as immunoreactivity to tuberculin (Tuberculin Sensitivity Test or TST) and presumably, the concomitant ‘trained immunity’ conferred by BCG vaccination at childbirth wanes rapidly within first few years of childhood in the absence of ‘booster’ *Mycobacterium spp*. exposures [REFs in 12,15,16], the prevalent actual ‘trained immunity’ and heterologous cell-mediated immunity correlate of any population could be a better evaluable predictive parameter of populations’ response to COVID-19 infections [5,12], if any, rather than relying on BCG vaccination at childbirth/childhood, the national universal vaccination policy, its coverage, or its implementation year as being proposed as well as explored [14,3,4]. The tuberculin immunoreactivity persistence is known to be associated with lower mortality from unrelated diseases both in infants as well as the elderly for a long time [REFs in 1-5,9-12,15].

Tuberculin Sensitivity Test (TST) and Interferon Gamma Release Assays (IGRAs) are used to indirectly assess the presence of memory T-cell response or cell-mediated immune response against previous *Mycobacterium spp*. antigens (environmental or BCG vaccine) exposure [15,17,18]. In the absence of clinically active tuberculosis disease, the presence of tuberculin immunoreactivity is referred to as ‘Latent Tuberculosis Infection’ (LTBI) broadly signifying the absence of tuberculosis disease but the presence of active immunity against the pathogen [17-19]. However, it should be noted that due to a lack of direct tests to ascertain the asymptomatic or Latent Tuberculosis infection (LTBI), the presence of immunoreactivity to mycobacterial antigens as determined by TST or IGRA in the absence of clinically active tuberculosis is currently defined as ‘LTBI’ by WHO for ‘being at-risk of developing TB’ as a part of ‘World Health Organization’s End TB strategy’ to identify/track the individuals at-risk for the management follow-up purposes [18,19]. It is acknowledged by the WHO that only a small fraction (5-10%) of these individuals may develop TB over the course of their lives [18,20]. Furthermore, it is estimated that upto 90% of LTBI individuals developing tuberculosis later could be attributed to reinfection on the waning of immunoreactivity (immunity) while remaining small minority could be attributed to actual reactivation of latent bacilli [17,19] on immunosuppression *(e.g*., HIV infection, cancer, immunosuppressant therapy) in genuinely LTBI individuals that may at the most could comprise 1-11 % of ‘LTBI’ labeled individuals in different settings as per an estimate based on several previous epidemiological studies [19]. This ‘LTBI’ nomenclature for persons at-risk of developing tuberculosis has been retained by WHO though not without causing confusion and general avoidance of its use as a proxy measure of imparted protective cell-mediated immunity or the associated ‘trained immunity’ of the population in the general parlance [19]. We reason, if indeed ‘trained immunity’ or persisting cell-mediated immunity conferred by exposure to *Mycobacterium spp*. (Environmental or BCG) could help reduce COVID-19 infections or mortality in a population, the estimated prevalence of the tuberculin immunoreactivity or the so-called ‘% LTBI’ [21] of resident populations would closely correlate with COVID-19 infection and mortality rates regardless of the BCG vaccination policy, BCG coverage or its implementation among countries [22]. The European countries that have a quite mix of diverse BCG vaccination policies – ranging from none ever to current universal vaccination [22], relatively comparable medical infrastructure, mobility, exposure to SARS-CoV-2, and other confounding variables and more importantly currently at a similar stage of epidemic-curve, *i.e*., post-infections-peak but could have differential ‘trained immunity’ and cell-mediated immunity status as may be supposed from %LTBI prevalence [5,6,7,12] offer an excellent opportunity to evaluate such an assertion.

## METHODS

The populations from 20 European countries with a differential prevalence of %LTBI [21] (published by the ‘Institute for Health Metrics and Evaluation (IHME)’ 2018) and comparable confounding variables, including the stage of the pandemic (infections peak) (Table 1) are assessed for any correlation with COVID-19 cases and mortality data of the ongoing pandemic from different stages, *i.e*., 8 April, 12 May, and 26 May 2020 from https://www.worldometers.info/coronavirus/ [23] without any exclusion criterion (e.g., age, sex, ethnicity) or data transformation/normalization, as done previously for vitamin D [24].

**Table 1:** COVID-19 (SARS-CoV-2) infections in European countries with different prevalence of Latent Tuberculosis Infection (% LTBI) and the Correlation Analysis

| SARS-COV-2 INFECTION (COVID-19) CASES & % LTBI IN EUROPEAN COUNTRIES |  |  |  |  |  |  |  |
| --- | --- | --- | --- | --- | --- | --- | --- |
|  | Most countries before infections peak |  | All countries post infections peak |  |  |  |  |
|  | 8 April 2020 |  | 12 May 2020 |  | 26 May 2020 |  |  |
| Countries | Cases per Million Pop. | Deaths per Million Pop. | Cases per Million Pop. | Deaths per Million Pop. | Cases per Million Pop. | Deaths per Million Pop. | % LTBI (2017) |
| Spain | 3137 | 314 | 5735 | 572 | 6060 | 580 | 6.06 |
| Iceland | 4736 | 18 | 5278 | 29 | 5290 | 29 | 7.67 |
| Ireland | 1230 | 48 | 4685 | 297 | 5015 | 327 | 8.14 |
| Belgium | 2019 | 193 | 4612 | 751 | 4959 | 806 | 8.75 |
| UK | 895 | 105 | 3286 | 472 | 3909 | 546 | 9.55 |
| Italy | 2306 | 292 | 3636 | 508 | 3813 | 545 | 12.87 |
| Switzerland | 2686 | 103 | 3506 | 213 | 3557 | 221 | 8.42 |
| Sweden | 834 | 68 | 2641 | 322 | 3412 | 409 | 10.07 |
| Portugal | 1289 | 37 | 2715 | 112 | 3040 | 132 | 10.33 |
| France | 1671 | 167 | 2718 | 408 | 2800 | 437 | 8.86 |
| Netherlands | 1199 | 131 | 2497 | 318 | 2661 | 342 | 8.29 |
| Germany | 1309 | 25 | 2060 | 91 | 2164 | 101 | 9.20 |
| Denmark | 933 | 38 | 1815 | 92 | 1974 | 97 | 8.81 |
| Turkey | 453 | 10 | 1657 | 46 | 1884 | 52 | 12.46 |
| Norway | 1123 | 19 | 1500 | 41 | 1547 | 43 | 8.46 |
| Estonia | 893 | 18 | 1312 | 46 | 1383 | 49 | 12.03 |
| Finland | 449 | 7 | 1080 | 49 | 1196 | 56 | 8.28 |
| Czechia | 488 | 9 | 763 | 26 | 845 | 30 | 11.41 |
| Hungary | 93 | 6 | 340 | 44 | 390 | 52 | 13.03 |
| Slovakia | 125 | 0.4 | 267 | 5 | 277 | 5 | 12.70 |
| Average | 1393.40 | 80.42 | 2605.15 | 222.10 | 2808.80 | 242.95 | 9.77 |
| STDEV | 1129.98 | 94.61 | 1600.99 | 221.41 | 1681.89 | 238.78 | 2.00 |
| CORRELATION ANALYSIS |  |  |  |  |  |  |  |
| Cases Per Million Pop. vs %LTBI | r(20): -0.5511<br>p – value: 0.0118 |  | r(20): -0.6338<br>p – value: 0.0027 |  | r(20): -0.6194<br>p – value: 0.0036 |  |  |
| Deaths Per Million Pop. vs %LTBI | r(20): - 0.2836;<br>p – value: 0.2256 |  | r(20): -0.3283;<br>p – value: 0.1576 |  | r(20): -0.3088;<br>p – value: 0.1853 |  |  |

*Note:* The values rounded off to the indicated decimal places. (Refer to main text and Supplementary file 1 for estimates about top four South American countries that had been severely hit by COVID-19, i.e., Brazil, Peru, Chile and Ecuador, along with the states formerly
belonging to East and West Germany. All display perfect negative covariation with LTBI prevalence.)

## RESULTS

The potential ‘trained immunity’ prevalence of the populations (i.e., %LTBI) display a strong correlation with COVID-19 infections (Table 1). Early-stage comparisons of the impact could be marred with changing local public policies and adherence to them (compare the change in COVID-19 incidence and mortality among countries in Table 1 over time; it could be also applicable to the analysis recently presented in reference [3 and 4]). Our analysis of the COVID-19 data reveals a consistently negative covariation of the cases per million with population’s %LTBI at all the time points evaluated [*r*(20): -0.5511 to -0.6338; p-value: 0.0118 to 0.0027] both pre- and post-infections peak [8 April to 26 May 2020], whereas the negative covariation of deaths per million population [r(20) – 0.2836 to – 0.3283] though improved post-infections peak did not reach statistical significance (p-values >0.05) (Table 1), similar to previously observed by us for other countries at an earlier phase of the pandemic [2]. See Figure 1 for the potential predictive correlative inference of the data on May 26, 2020.

**Figure 1.**
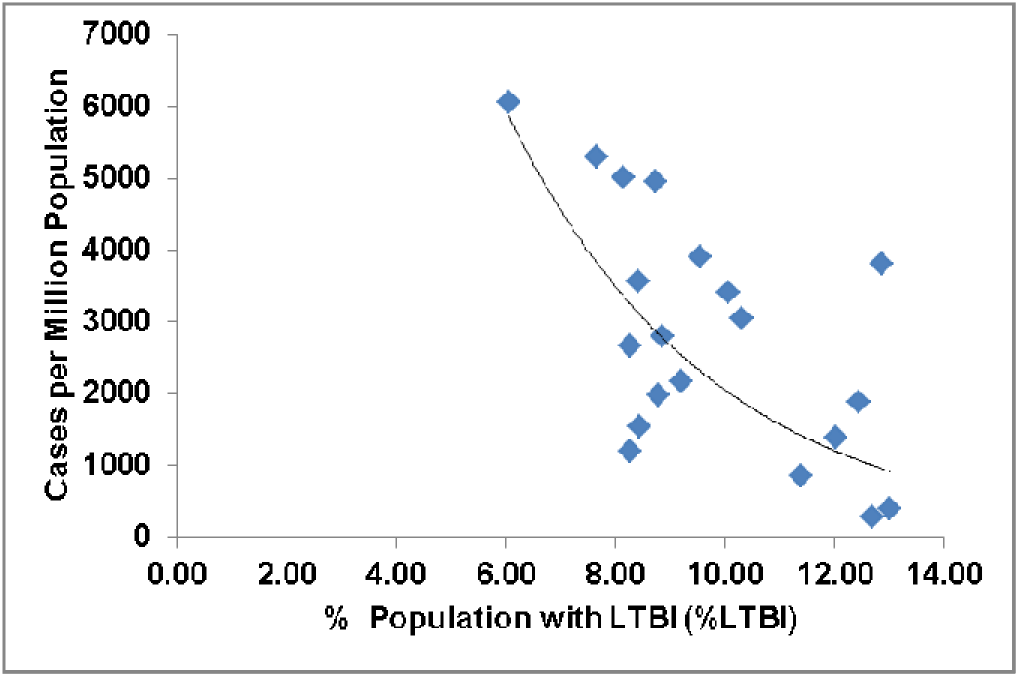
The prevalence of %LTBI in European countries negatively correlated with COVID-19 cases per million population (26 May 2020)

## DISCUSSION

In the differential tuberculin immunoreactive populations of European countries, the COVID-19 cases per million population negatively covary with the %LTBI prevalence at each analyzed stage of the pandemic. However, this negative correlation observed with deaths per million populations does not reach statistical significance as could be partly expected from individuals/population components without sufficient protective ‘trained immunity’ or cell-mediated immunity being prone to getting infected and the non-adjustment of other underlying confounding variables including but not limited to age, medical practices, and medical infrastructure and support. The transformation of data and its adjustment for confounding variables as done in the recent reports [3, 4] are expected to further improve the correlation observed in the current study but its relevance cannot be ensured rather we should allow space for other potential protective variables to be discovered, discussed and evaluated. In short, regardless of the childhood BCG vaccination status, the cell-mediated tuberculin immunoreactivity or ‘LTBI’ prevalence could be argued to be more important in assessing the functional potentially protective trained immunity and cell-mediated heterologous immunity of a population.

The strength of the assertion lies in the large affected study population, *i.e*., 1,413,367 COVID-19 patients (about 25% of total worldwide cases on 26.05.2020) [23]; representing 20 different %LTBI prevalence groups/populations from countries ranging from never ever to current mandatory BCG vaccination policy; comparable health infrastructure, screening and reporting guidelines along with similar age and sex distribution and other confounding variables. The correlation analysis consistently indicates the same association at all 3 different time points of the epidemic curve evaluated indicating a potential protective impact of the populations’ immunoreactivity to mycobacterial antigens regardless of source (BCG or environmental mycobacteria). Thus, the LTBI prevalence in countries could very well satisfactorily explain the COVID-19 impact regardless of the consideration of BCG vaccination policy or its implementation [3,4,14].

The states belonging to the former East and West Germany present a peculiar case for putting the current assertions to test with respect to other recently reported studies that find a protective role for BCG vaccination policy on COVID-19 incidences and mortality. Currently, formal estimates of LTBI prevalence for East and West Germany are not available [21]. However, as per a tuberculosis contact tracing study performed in a police academy in 2006 by Diel et al. the estimated differential TST positivity in personnel (age group 15-62 years) from East and West Germany were 27% LTBI (89.2 % BCG vaccinated) and 16 % LTBI (30.7% BCG vaccinated), respectively [25]. Assuming the sample to be representative of the population, the 8% annual decrease in TST positivity as expected from previous estimates for adults [16] would predict West Germans to have already reached the average background level of 9.2 %LTBI for Germany in 2017 while the East Germans maybe still having higher cell-mediated immunity and trained immunity as could be expected from estimated prevailing LTBI of 22.5 % for the same age-progressed group (now 30-78 years) (Supplementary File 1). The observed average COVID-19 incidence in East Germany (102.99 per million) had consistently remained lower than West Germany (214.55 per million) as per the estimate available on 22 May 2020 available at https://www.citypopulation.de/en/germany/covid/ (excluding the city-states Berlin, Bermen and Hamburg) [On 17 Aug 2020 the incidences were: East Germany 121.74 per million; West Germany 262 per million] (Supplementary File 1). Similarly, many fold lower mortality has been consistently observed in East Germany till now as per the available data at https://www.citypopulation.de/en/germany/covid/ despite having higher elderly population [3]. The countries in southern hemisphere Australia and New Zealand with the comparable prevalence of LTBI (Australia: 10.12%; New Zealand: 11.36%), supposedly with comparable confounding variables (e.g., social and medical practices, access to health services, infrastructure, population composition, age distribution, general health, etc) to European countries and at the similar stage of pandemic (already passed first peak of infections), have experienced quite a benign impact of COVID-19 (Australia: 280 cases and 4 deaths per million; New Zealand: 312 cases and 4 deaths per million as on 26 May 2020) as has been observed for higher LTBI prevalence European country Slovakia (12.7% LTBI; 277 cases and 5 deaths per million as on 26 May 2020)[23]. There may be other potentially protective variables at play as well that may be contributing to this reduced incidence and mortality such as vitamin D levels, Zinc levels, etc which may be beyond the scope of current discussion [24,26]. Strikingly, similar negative covariation of COVID-19 incidence and mortality with estimated ‘LTBI’ prevalence can be seen in the groups of other countries/populations which can be assumed to have comparable confounders and at a similar stage of the pandemic for the purpose of comparison, *e.g*., the incidence of COVID-19 per million population in four top worst-affected countries of South America (>1500 cases/million population) namely, Brazil (1847 cases per million; %LTBI: 26.05), Ecuador (2121 cases per million; 18.2% LTBI), Peru (3941 cases per million; %LTBI:17.32), and Chile (4082 cases per million; %LTBI: 13.79) increased with decreasing % LTBI (perfect negative covariation) independent of their BCG vaccination policy or coverage as on 26 May 2020 [23] (see Supplementary File 1). Among them Ecuador does not have current vaccination policy while it had it in the past, the other three nations have universal BCG vaccination policy in which Chile reports the most wider coverage. However, it should be noted that these comparisons/covariations would supposedly start to become reliable/stable once at least all countries with supposed similar confounders, have passed the peak of infections [24] and could progressively reach the maximum level of reliance at the end of current pandemic – when outcome of each infection becomes known.

Multiple past studies indicate that the presence of tuberculin reactivity, not the BCG vaccination history in the young as well as the old to be negatively correlated with the incidence of several diseases including respiratory diseases [REFs in 1,3-12,15] and the elderly had been recommended to remain TST positive to reduce chances of unrelated disease and pneumonia [9,10]. It must be remembered that BCG vaccination does not necessarily elicit a cell-mediated immune response in all, at the same rate and there is also reported loss with age or absence of re-challenge and immune suppression. In the view of observation presented, it may be suggested that ongoing trials/studies evaluating the effect of BCG vaccination on COVID-19 infections [1,13], including the one recently concluded in Israel and reported in JAMA [14] could provide more objective conclusions on inclusions of the estimates about the ‘trained immunity’ and heterologous cell-mediated immune response of study participants/ populations [5,12].

It is pertinent to mention here that the statistical correlations no matter how significant they appear from covariation/correlation value, and the associated p-value significance levels, they never indicate a cause and effect relationship. Till the cause and effect relationships are unequivocally established for variables based on evidence, efforts to make predictions/projections and overfitting of the data may be better avoided – giving a false sense of cause and effect relationship. Statistical correlation of COVID-19 with BCG vaccination policy, coverage, or implementation year that is being presented in the literature using highly transformed variables that give high R^2^ value and low p-values and being hotly discussed, even if it had been proven, are equally of no interventional use in the current scenario as there is no means to go in the past and vaccinate the current vulnerable population of the elderly and people with comorbidities nor change GDP, migration rate, population density, etc. [REFs in 2-4]. Rather resources may be channelized to evaluate/ explore the potentially protective co-varying variables that possibly may have cause and effect relationship – employing appropriate controls, and would be amenable to intervention, no matter how weak the relationship may appear.

Dedicated studies using the available patient records or epidemiological surveys backed by follow-up clinical trials with suitable controls for the ‘trained immunity’ and heterologous cell-mediated immunity correlates are advisable to decisively assess the biological significance/evidence of the current and previously observed correlations or their absence to put the ongoing debate to rest and reach at a more meaningful conclusion about the interventional usage of non-specific trained immunity and heterologous cell-mediated immunity in COVID-19 control.

## Data Availability

All data referred in the manuscript are included in the manuscript.

## Acknowledgment

The general funding support by Banaras Hindu University to the laboratory of SS is acknowledged. SS is a Ramalingaswami Fellow, DBT, India (BT/RLF/Re-Entry/50/2011).

## Compliance with ethical standards

### Ethical statement

The study complied with the existing ethical standards.

---

**Supplementary File 1: Sheet 1 – E. and W. Germany States:** The incidence of COVID-19 in East and West Germany states starting 10 April 2020 to 17 Aug 2020 provided. The incidence of in COVID-19 in East Germany (22.5% LTBI estimated) consistently remained lower than West Germany (9.2 % LTBI). (Estimated LTBI is for 30-78 yr age group). **Sheet 2 – South America COVID-19:** The incidence among four top COVID-19 affected countries Brazil, Ecuador, Peru and Chile negatively covaries with %LTBI prevalence [Note perfect covariation observed on later stage (26 May 2020) as compared to earlier stage (12 May 2020) of pandemic as expected].

